# Feasibility and results of joint ambulatory monitoring of exposure to particulate matter pollution and lung function in children in Abidjan, Côte d’Ivoire, a cross-sectional observational study

**DOI:** 10.1101/2025.08.16.25333819

**Authors:** Auriane Pajot, Marie Yapo, Cathy Liousse, Madina Doumbia, Sylvain Gnamien, Stéphane Ahoua, Sonia Dje, Flore Dick Amon Tanoh, Michael Fayon, Véronique Yoboue, Olivier Marcy

## Abstract

**Background:** Children are largely exposed to air pollution in low- and-middle income countries, yet data on exposure and respiratory effects of air pollution remain limited. This study aimed to assessed the feasibility and outcomes of joint ambulatory monitoring of exposure to fine particulate matter (PM_2_._5_) and spirometry in children living in Abidjan, Côte d’Ivoire.

**Methods:** We did a cross-sectional observational study among children aged 7-17 years. After a baseline spirometry, children were asked to wear portable PM_2.5_ sensors and to perform 2×3 daily flow-volume curves using a portable spirometer for 7 days. We described the proportion of acceptable measurements, percent predicted forced expiratory volume (ppFEV1), and hourly geometric mean PM_2.5_ concentrations, and analyzed the cumulative delayed effects of PM_2_._5_ on ppFEV1 using distributed lag non-linear models (DLNMs).

**Results:** Of 29 children enrolled - 18 (62.1%) female, median age 12 years - all performed spirometry with 1101 (90.4%) of 1218 expected flow-volume curves obtained; 313 (28.4%) valid non-duplicate curves were analyzed. The median ppFEV_1_ was 79.6% [71.5-87.4]; with lower values in the morning than in the evening (p < 0.001). Of 146,160 expected PM_2.5_ measurements, 93,689 (64.1%) were obtained; 6,328 aberrant data were excluded. The median hourly PM_2.5_ concentrations were 164.2 [107.0 - 272.2] µg/m^3^. PM_2.5_ levels varied throughout the day with pollution peaks observed in the morning. A significant decrease in ppFEV_1_ was observed between 0 and 2 hours post-exposure, after an interquartile range increase of 120.9 µg/m^3^ in PM_2.5_ exposure (β = −2.21; CI [−3.74; −0.69]).

**Conclusion:** Ambulatory spirometry and PM_2.5_ measurements are feasible with portable devices in African children. High PM_2.5_ exposure and individual variability in lung function highlight the need for further research respiratory effects of air pollution in children.

**Strengths and limitations of this study:**

- Innovative combination of repeated ambulatory spirometry and personal PM_2_._5_ monitoring allowing to capture individual level, real-time exposures and lung function in children from an under researched setting over 7 days.
- High proportion of collected spirometry curves and PM_2_._5_ data demonstrates the feasibility and acceptability of mobile monitoring tools among children living in precarious urban environments.
- Relatively small sample size, short follow-up period, and absence of environmental covariates restrict the generalizability of our findings.
- Study providing novel evidence of short-term respiratory effects of extreme PM_2_._5_ exposures in children, underscoring the importance of incorporating mobile, individual-based approaches into future research and public health interventions.
- Study underscoring the importance of incorporating mobile, individual-based approaches into future research and public health interventions.

## INTRODUCTION

Air pollution, and particularly exposure to fine particles with a diameter of less than 2.5 microns (PM_2.5_), is a well-established risk factor for respiratory diseases. Due to their small aerodynamic size, PM_2_._5_ particles can penetrate deep into the respiratory tract, reaching the alveoli and even entering the bloodstream, where they may trigger acute and chronic inflammatory responses (1,2). Children are especially vulnerable to particulate pollution due to physiological specificities compared to adults including a higher respiratory rate, a still developing pulmonary system, and increased proximity to the ground where particulate matter concentrations are higher (3). Numerous studies conducted in high-income countries have demonstrated associations between exposure to PM_10_ and PM_2_._5_ and adverse respiratory outcomes in children, including reduced lung function (4–9) and increased risk of asthma onset and exacerbation (10–12). In contrast, far less evidence is available from low- and middle-income countries (LMICs), where exposure to air pollution is often more intense due to widespread biomass combustion for cooking and food preservation (13). In such settings, young children may be exposed for extended periods of time, either within the household or accompanying mothers during polluting activities. However, exposure levels and their respiratory consequences remain poorly characterized. A pilot study conducted in Abidjan, Côte d’Ivoire, reported median PM_2_._5_ concentrations reaching 60.8 μg/m^3^ indoors and 58.2 μg/m^3^ in residential outdoor areas (14), about four times the WHO’s 24-hour guideline.

Few studies have assessed the impact of personal exposure to PM_2.5_ on the respiratory function of children by combining the use of spirometers and portable PM sensors PM_2.5_ (5,8,10,15). Although the use of portable sensors and spirometers has been validated in adults (16), their use in children poses challenges in terms of understanding and adherence, particularly without medical supervision (17,18). These constraints are even more pronounced in disadvantaged areas, where educational resources and parental support may be limited (19,20). There is therefore a critical need to assess the feasibility of combining personal pollution monitoring and self-administered spirometry in children living in vulnerable environments.

This study aimed to evaluate the feasibility and outcomes of using portable PM_2_._5_ monitors together with ambulatory spirometry in children living in urban disadvantaged areas of Abidjan. We examined variations in exposure and respiratory health across three groups of children with differing levels of biomass combustion exposure. We hypothesized that higher personal PM_2_._5_ exposure would be associated with short-term reductions in lung function, particularly in forced expiratory volume in one second (FEV_1_).

## METHODS

### Study design and population

We conducted a cross-sectional observational study from February 2023 to February 2024 involving 30 children aged 7 to 17 years. Participants were selected from a sub-cohort of the APIMAMA Kids study, a pediatric extension of the APIMAMA project (2022–2026, https://apimama.org), which investigates domestic air pollution from biomass combustion and its respiratory effects on women and children in Côte d’Ivoire.

Participants were drawn from three groups categorized by the primary source of biomass combustion exposure associated with maternal domestic or occupational activities: (G1) children whose mothers use charcoal for cooking, (G2) children whose mothers smoke fish as a livelihood, and (G3), children whose mothers primarily use butane gas for cooking. After informed consent was signed by the legal representative and assent was obtained from the children, those children included in APIMAMA Kids who were able to understand and express themselves in French. It was proposed that children independently perform flow-volume curves using a Spirotel (Mir France) as part of the ambulatory measurements.

Each participant wore a personal air pollution monitor to assess real-time PM_2_._5_ exposure and performed multiple daily lung function tests using a handheld spirometer over a continuous 7-day period. Ten sets of portable spirometers and pollution sensors were rotated across participants in three successive field campaigns, with children assigned according to their group.

### Study settings

All participants lived in urban areas in Yopougon, the most densely populated district of Abidjan with nearly 2 million inhabitants spread over three distinct neighborhoods (21). Each study group was recruited from a distinct area of Yopougon, selected based on predominant types of household or occupational biomass use. Data collection took place in participants’ residential environments under ambulatory (non-clinical) conditions. Fieldwork was conducted in three successive phases: northern Yao Séhi (G1), southern Yao Séhi (G3), Yopougon Santé and Abobodoumé (G2).

### Study procedures

Mothers completed questionnaires regarding household characteristics, living conditions, and their child’s respiratory symptoms, based on the standardized ISAAC questionnaire. A clinical examination and baseline spirometry, including a bronchodilator reversibility test, were performed by a qualified healthcare professional using a spirometer (Spirolab, MIR France). These initial flow–volume curves served as a reference to assess the quality of independently performed spirometry by the children. Spirometry was conducted and interpreted according to the Global Lung Function Initiative reference equations (GLI 2012) and in line with ATS/ERS technical standards, with adaptations appropriate for pediatric populations (22). The spirometric parameters collected included forced vital capacity (FVC), forced expiratory volume in one second (FEV_1_), the FEV_1_/FVC ratio (Tiffeneau’s index), and forced expiratory flow at 25–75% of the pulmonary volume (FEF_25-75_).

Following baseline assessment, children received a portable spirometer (Spirotel, Mir France) pre-calibrated with 2012 GLI parameters. They were trained to perform the maneuver independently, instructed to: sit or stand comfortably, insert a disposable mouthpiece, apply a nose clip, take a maximal inhalation, and then exhale forcefully into the turbine-connected mouthpiece. Two consecutive spirometry curves were requested three times per day, in the morning (until 11:30 a.m.), in the middle of the day (11:30 a.m.–5:30 p.m.) and in the evening (5:30 p.m.–midnight), for a total of six curves per day for one week.

Each child also received a portable low-cost optical particle sensor (GAIA APIMAMA, Plantower PMS5003) designed for the APIMAMA project (23), which measured PM_2_._5_ concentrations every two minutes. Children were instructed to wear the sensor across their chest throughout the day, excluding intense physical activity and to charge it overnight in their bedroom to resume use the following morning. Twice during the measurement period, trained field technicians retrieved and downloaded data from the sensors to avoid data loss due to memory saturation.

Spirometry data were extracted using the Winspiro PRO software at the end of the collection period. Ambulatory spirometry curves were retained according to the ATS/ERS (American Thoracic Society / European Respiratory Society) visual acceptability criteria, defined by a maximal effort, a rapid start without hesitation, a complete expiration free of artifacts, and the achievement of an expiratory plateau. When multiple acceptable curves were recorded for the same time period, only the highest predicted FEV_1_ percentage (ppFEV_1_) was retained, limiting inclusion to one curve per time window per day (24). All health data were entered digitally into REDCap and integrated into a dedicated research database. PM_2_._5_ values were calibrated using correction factors specific to each group: 5.5 for Groups 1 and 3, and 5.0 for Group 2 (25). Concentrations below 6 µg/m^3^, corresponding to the sensor’s detection threshold, were excluded. Hourly and daily averages were calculated for each participant.

### Outcomes and variables of interest

Feasibility outcomes included the frequency and quality of flow-volume curves independently performed by the children and the proportion of valid individual PM_2.5_ concentration data collected throughout the study period. The main outcomes were individual PM_2.5_ exposure measurements and intra-individual variations in ppFEV_1_ collected by portable spirometry.

Key independent variables comprised average PM_2_._5_ concentrations calculated as hourly means and moving averages over 1-hour, 8-hour, and 24-hour intervals, along with time of day (morning, midday, evening). Covariates included child’s age, sex, baseline asthma status (based on initial spirometry and clinical questionnaire), study group (reflecting different biomass combustion exposure levels), and school attendance status (attending vs. not attending school). These were included as adjustment variables in multivariable models.

### Data analyses

Descriptive statistics were used to summarize the number and validity of spirometry recordings, the proportion of usable PM_2_._5_ measurements, and the demographic and respiratory characteristics of participants.

We assessed associations between ppFEV_1_ and PM_2_._5_ using mixed-effects models for repeated measures, including linear mixed-effects models (repeated-measures ANOVA) and generalized linear mixed-effects models (GLMM). Post-hoc tests were performed for categorical variables >2 levels when p <0.05. Short-term lagged effects of PM_2_._5_ exposure on lung function were evaluated using distributed lag models (DLMs), based on 24-hour and peak 1-hour and 8-hour moving averages.

To assess cumulative delayed effects up to 48 hours prior to FEV_1_ measurement, we applied distributed lag non-linear models (DLNMs) with mixed effects including a fifth-degree polynomial lag function and an AR(1) structure to account for temporal correlation (5). Analyses were conducted for the full cohort and stratified by time of day and exposure group. Results were reported as percent change in ppFEV_1_ per interquartile range (IQR) increase in 24-hour moving average PM_2_._5_.

Models were adjusted for confounders. Missing data were linearly interpolated (for gaps ≤ 6 hours), and observations were retained only when at least 50% of hourly PM_2_._5_ data were available for the 24-hour window preceding spirometry.

Analyses were performed using R, with significance set at p < 0.05, interaction terms with p < 0.10 indicated potential effect modification.

### Ethical aspects

The study protocol was approved (N/Réf : 177-23/MSHPCMU/CNESVS-km) by the National Ethics Committee for Life Sciences and Health in Côte d’Ivoire (CNESVS). In addition, authorizations were obtained from local chiefs to conduct the study in each neighborhood.

### Patient and public involvement

Children and their mothers contributed to optimizing sensor and spirometry placement and provided feedback on equipment acceptability. They also reported any issues during data collection, allowing prompt intervention. Results will be communicated in a format accessible to both children and adults.

## RESULTS

We included 29 children with a median age of 12 [11–14] years, 18 (62.1%) of whom were female, evenly distributed across the 3 groups (Table 1). Of the 30 children initially planned, one participant could not be included in group 2, as no additional child met the inclusion criteria for this group. Noisy breathing or chest wheezing at any point in life was reported by 5 (17.2%) of participants, with 2 (6.9%) experiencing wheezing in the past 12 months and 7 (24.1%) reported dry nocturnal cough in the past 12 months. No significant differences were observed between the 3 groups in terms of demographic, anthropometric and ISAAC characteristics. However, ppFVC was higher in G2 (100.6%) compared to G1 (78.1%) and G3 (86.4%) (p = 0.008). A total of 9 children (31.0%) had a ppFVC below 80% with 6 (60.0%) in group G1, 1 (11.1%) in G2, and 2 (20.0%) in G3 (p = 0.046). Regarding ppFEV_1_, 6 (20.7%) had values below 80%, including 4 in G1, 1 in G2 and G3 (p = 0.176) (Table 1).

**Table 1.** Baseline demographic, anthropometric, and spirometric characteristics of study participants.

| Characteristic | Overall<br>N = 29 | G1<br>N = 10 | G2<br>N = 9 | G3<br>N = 10 | p-value * |
| --- | --- | --- | --- | --- | --- |
| Age (years) | 12 [11-14] | 12.5 [11-13.75] | 12 [12-13] | 11.5 [11-14] | 0.880 |
| Gender female | 18 (62.1) | 8 (80.0) | 5 (55.6) | 5 (50.0) | 0.342 |
| Weight (kg) | 38.0 [29.0-45.0] | 40.0 [30.3-44.5] | 38.0 [29.0-47.0] | 32.5 [30.0-41.8] | 0.908 |
| Height (cm) | 145.0 [137.0-153.0] | 146.5 [140.3-151.0] | 145.0 [137.0-154.0] | 142.0 [137.5-151.5] | 0.870 |
| BMI | 17.1 [15.1-19.0] | 17.0 [15.1-19.4] | 17.9 [15.5-19.0] | 15.8 [15.2-18.7] | 0.930 |
| Schoolchildren | 26 (89.6) | 10 (100.0) | 8 (88.9) | 8 (80.0) | 0.339 |
| ISAAC questionnaire (Asthma) |  |  |  |  |  |
| Noisy breathing/whistling in the chest in the past | 5 (17.2) | 2 (20.0) | 1 (11.1) | 2 (20.0) | 0.842 |
| Noisy breathing/whistling in the chest (last 12 months) | 2 (6.9) | 1 (10.0) | 1 (11.0) | 0 | 0.754 |
| 1 to 3 wheezing attacks (last 12 months), N*=2 | 2 (100) | 1 (10.0) | 1 (11.0) | 0 | - |
| Sleep disturbed by wheezing (last 12 months), N*=15 |  |  |  |  | 0.999 |
| Less than one night a week | 1 (50.0) | 0 | 1 (100.0) | 0 |  |
| One or more nights a week | 1(50.0) | 1(100.0) | 0 | 0 |  |
| Speech limited to one or two words (last 12 months) | 2 (100.0) | 1 (100.0) | 1 (100.00) | 0 | — |
| Wheezing in the chest during or after exercise (last 12 months) | 4 (13.8) | 2 (20.0) | 1 (11.1) | 1 (10.0) | 0.779 |
| Dry cough at night (last 12 months) | 7 (24.1) | 4 (40.0) | 1 (11.1) | 2 (20.0) | 0.316 |
| Asthma diagnosed | 2 (6.9) | 1 (10.0) | 0 | 1 (10.0) | 0.999 |
| Baseline spirometric characteristics |  |  |  |  |  |
| FVC (mL) | 1.98 [1.62-2.32] | 1.96 [1.35-2.03] | 2.28 [1.93-2.89] | 2.01 [1.69-2.28] | 0.312 |
| ppFVC | 87.27 [79.56-96.57] | 78.06 [66.53-81.88] | 100.62 [88.53-105.53] | 86.4 [84.98-93.55] | **0.008** |
| ppFVC <80% | 9 (31.0) | 6 (60.0) | 1 (11.1) | 2 (20.0) | **0.046** |
| FEV1 (mL) | 1.91 [1.48-2.06] | 1.9 [1.35-1.97] | 1.95 [1.79-2.42] | 1.83 [1.52-1.98] | 0.651 |
| ppFEV<sub>1</sub> | 91.32 [85.0-101.41] | 86.33 [73.61-89.76] | 101.41 [91.79-106.61] | 91.33 [85.87-95.51] | 0.109 |
| ppFEV<sub>1</sub> < 80% | 6 (20.7) | 4 (40.0) | 1 (10.0) | 1 (10.0) | 0.176 |
| FEV1/FVC | 94.9 [90.6-99.5] | 99.5 [97.28-100] | 90.6 [87.9-92.7] | 93.1 [90.65-96.4] | 0.003 |
| ppFEV1/FVC | 106.01 [100.94-110.29] | 110.32 [107.91-111.28] | 100.94 [98.17-103.24] | 104.19 [100.78-108.57] | 0.006 |
| ppFEV1/FVC < 80% | 2 (6.9) | 0 | 1 (11.1) | 1 (10.0) | 0.566 |
| PEF (mL) | 3.61 [3.11-4.32] | 3.78 [3.01-4.54] | 3.5 [2.74-3.74] | 3.89 [3.34-3.99] | 0.647 |
| ppPEF | 77.3 [56.03-93.81] | 76.77 [54.93-96.47] | 66.14 [57.68-98.93] | 81.81 [59.28-89.66] | 0.990 |
| ppPEF <80% | 15 (51.7) | 6 (60.0) | 5 (55.6) | 4 (40.0) | 0.645 |
| FEF25-75 (mL) | 2.4 [1.94-2.76] | 2.55 [2.34-2.69] | 2.22 [1.68-3.2] | 2.19 [1.9-2.74] | 0.624 |
| ppFEF25-75 | 92.93 [84.35-113.79] | 91.42 [85.48-106.49] | 96.1 [85.06-113.79] | 90.17 [81.17-115.73] | 0.982 |
| ppFEF25-75 < 70% | 3 (10.3) | 0 | 2 (22.2) | 1 (10) | 0.283 |
| Reversible FEV<sub>1</sub> on baseline spirometry (presomptive asthma) | 4 (13.8) | 2 (20.0) | 1 (11.1) | 1 (10.0) | 0.779 |

n (%); Median [IQR] \*Fisher's exact test; Kruskal-Wallis rank sum test; Chi2 test
Measured parameters: forced vital capacity (FVC), forced expiratory volume in 1 second (FEV<sub>1</sub>), forced expiratory flow at 25–75% of FVC (FEF25-75), and FEV1/FVC ratio. Reversible airway obstruction (FEV<sub>1</sub> increase >12% post-bronchodilator) was used as a presumptive diagnosis of asthma. (G1) children of mothers who use charcoal for cooking, (G2) children of mothers who smoke fish, and (G3) children of mothers who use butane gas for cooking.

A total of 1,101 (90.4%) flow-volume curves were obtained out of 1,218 expected (Figure1). The median number of curves performed per child was 36 [29–42] over 7 days. Boys performed more curves than girls, with a median of 42 [36.5–59] versus 33 [27.2–36], (p = 0.005) (Appendix 1).

**Figure 1.**
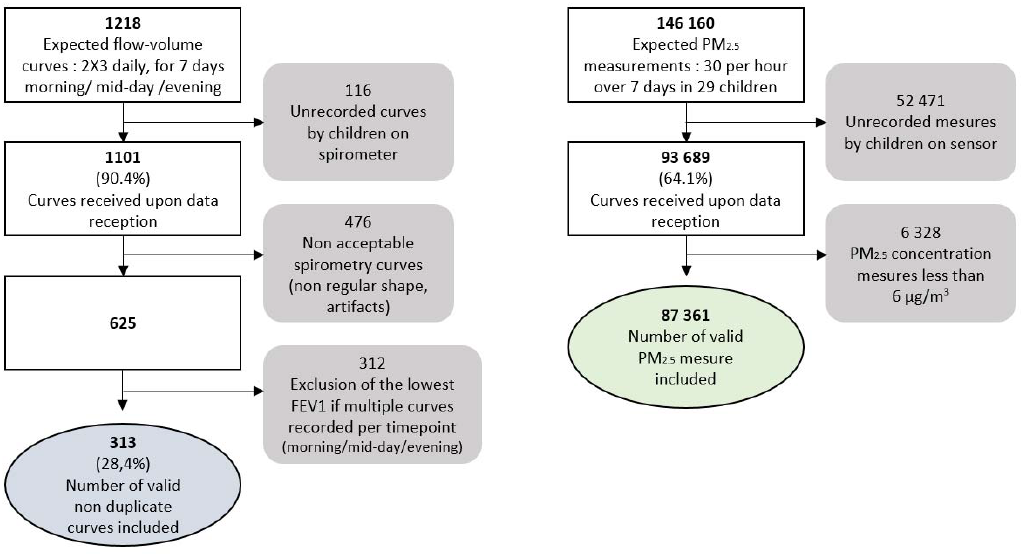
Flow chart of data selection from spirometry curves and PM_2.5_ concentration measurements in 29 children

Of the 1,101 curves collected, 476 were excluded due to non-compliance with the ATS/ERS validation standards. Ultimately, 313 (28.4%) valid, non-duplicate curves were included in the analyses (Figure1).

Of the 313 valid spirometry measurements, the median FVC and FEV_1_ values were 1.82 L [1.54-2.09] and 1.72 L [1.5-1.9], respectively. 106 (33.9%) had a ppFEV_1_ below 80%, with higher proportions in G1 (56, 44.1%) compared to G2 and G3 (11, 16.4% and 39, 32.8%, respectively; p = 0.228) and 161 (51.4%) had a ppFVC below 80% with a significantly higher proportion in G1 with 89 (70.1%) compared to G2 and G3 (22,32.8% and 50, 42.0%; p = 0.047) (Table 2).

**Table 2.** Characteristics of valid ambulatory spirometry measurements.

|  | Overall | G1 | G2 | G3 |  |
| --- | --- | --- | --- | --- | --- |
| Spirometry | N =313 | N =127 | N = 67 | N = 119 | p.value* |
| FVC (mL) | 1.82 [1.54-2.09] | 1.82 [1.54-2.08] | 1.81 [1.6-2.4] | 1.83 [1.52-2.04] | 0.503 |
| ppFVC | 79.62 [71.53-87.43] | 75.18 [66.85-81.46] | 84.31 [75.39-95.18] | 82.2 [73.59-87.9] | 0.057 |
| ppFVC <80% | 161 (51.4) | 89 (70.1) | 22 (32.8) | 50 (42.0) | 0.047 |
| FEV1 (mL) | 1.72 [1.5-1.96] | 1.8 [1.54-2] | 1.71 [1.52-2.26] | 1.68 [1.42-1.82] | 0.760 |
| ppFEV1 | 85.31 [76.74-93.58] | 82.3 [73.88-90.45] | 90.4 [82.02-100] | 86.19 [76.47-92.54] | 0.221 |
| ppFEV1 < 80% | 106 (33.9) | 56 (44.1) | 11 (16.4) | 39 (32.8) | 0.228 |
| FEV1/FVC | 98.5 [93.2-100] | 100 [98.8-100] | 97.8 [91.45-100] | 94.4 [89.75-98.85] | 0.097 |
| ppFEV1/FVC | 109.96 [104.17-111.62] | 110.2 [109.77-111.62] | 109.53 [101.43-113.67] | 105.31 [100.46-110.41] | 0.212 |
| ppFEV1/FVC < 80% | 9 (2.9) | 2 (1.6) | 4 (6.0) | 3 (2.5) | 0.671 |
| PEF (mL) | 3.73 [3.09-4.57] | 3.85 [3.18-4.84] | 3.6 [2.88-4.26] | 3.76 [3.02-4.39] | 0.641 |
| ppPEF | 73.43 [62.38-89.32] | 80.53 [62.64-100.83] | 70.64 [62.05-81.2] | 71.46 [62-83.47] | 0.596 |
| ppPEF <80% | 196 (62.6) | 61 (48.0) | 48 (71.6) | 87 (73.1) | 0.302 |
| FEF25-75 (mL) | 2.35 [1.95-2.93] | 2.67 [2.24-3.09] | 2.35 [1.69-2.66] | 2.15 [1.77-2.61] | 0.376 |
| ppFEF25-75 | 97.07 [79.02-115.48] | 106.69 [83.51-131.93] | 105.1 [81.78-117.22] | 92.34 [75.82-103.61] | 0.380 |
| ppFEF25-75 < 70% | 56 (17.9) | 18 (14.2) | 12 (17.9) | 26 (21.8) | 0.812 |

n (%); Median [IQR] \* Repeated-measures ANOVA (linear mixed-effects model), generalized linear mixed-effects model with repeated measures
(G1) children of mothers who use charcoal for cooking, (G2) children of mothers who smoke fish, and (G3) children of mothers who use butane gas for cooking

The ppFEV_1_ measured in the evening was significantly higher than that measured in the morning or midday, with a median of 86.1 [75.3 – 93.4]%, 85.9 [79.3 – 94.1]%, and 82.35 82.5 [75.8 – 92.1]%, respectively (p < 0.001). Median ppFEV_1_ measurements in non-asthmatic children (n = 267) were higher (87.2 [79.0–94.5]%) compared to asthmatic children (72.4 [67.0–82.1]%) with a borderline significant difference (p *=* 0.052) (Table 3).

**Table 3.**
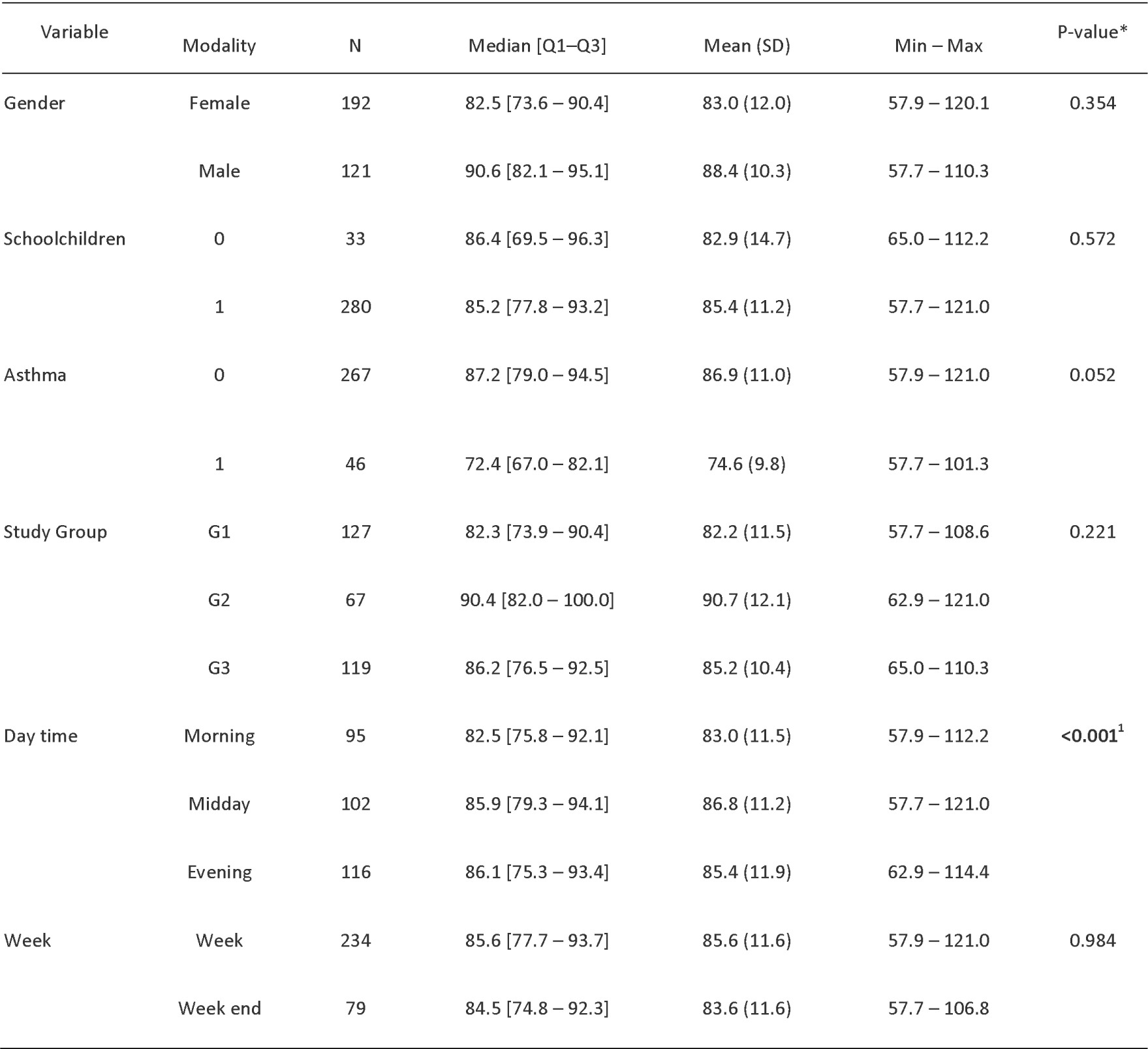
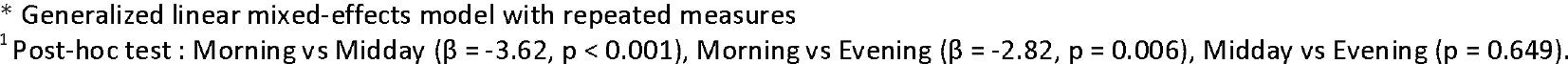
ppFEV_1_ variations according to children’s characteristics and timing.

| Variable | Modality | N | Median [Q1–Q3] | Mean (SD) | Min – Max | P-value* |
| --- | --- | --- | --- | --- | --- | --- |
| Gender | Female | 192 | 82.5 [73.6 – 90.4] | 83.0 (12.0) | 57.9 – 120.1 | 0.354 |
|  | Male | 121 | 90.6 [82.1 – 95.1] | 88.4 (10.3) | 57.7 – 110.3 |  |
| Schoolchildren | 0 | 33 | 86.4 [69.5 – 96.3] | 82.9 (14.7) | 65.0 – 112.2 | 0.572 |
|  | 1 | 280 | 85.2 [77.8 – 93.2] | 85.4 (11.2) | 57.7 – 121.0 |  |
| Asthma | 0 | 267 | 87.2 [79.0 – 94.5] | 86.9 (11.0) | 57.9 – 121.0 | 0.052 |
|  | 1 | 46 | 72.4 [67.0 – 82.1] | 74.6 (9.8) | 57.7 – 101.3 |  |
| Study Group | G1 | 127 | 82.3 [73.9 – 90.4] | 82.2 (11.5) | 57.7 – 108.6 | 0.221 |
|  | G2 | 67 | 90.4 [82.0 – 100.0] | 90.7 (12.1) | 62.9 – 121.0 |  |
|  | G3 | 119 | 86.2 [76.5 – 92.5] | 85.2 (10.4) | 65.0 – 110.3 |  |
| Day time | Morning | 95 | 82.5 [75.8 – 92.1] | 83.0 (11.5) | 57.9 – 112.2 | <0.001<sup>1</sup> |
|  | Midday | 102 | 85.9 [79.3 – 94.1] | 86.8 (11.2) | 57.7 – 121.0 |  |
|  | Evening | 116 | 86.1 [75.3 – 93.4] | 85.4 (11.9) | 62.9 – 114.4 |  |
| Week | Week | 234 | 85.6 [77.7 – 93.7] | 85.6 (11.6) | 57.9 – 121.0 | 0.984 |
|  | Week end | 79 | 84.5 [74.8 – 92.3] | 83.6 (11.6) | 57.7 – 106.8 |  |

\* Generalized linear mixed-effects model with repeated measures
<sup>1</sup> Post-hoc test : Morning vs Midday ( $\beta = -3.62$ , $p < 0.001$ ), Morning vs Evening ( $\beta = -2.82$ , $p = 0.006$ ), Midday vs Evening ( $p = 0.649$ ).

Of the 146,160 PM_2.5_ measurements expected for 29 children (30 per hour over 7 days per child), 93,689 (64.1%) were obtained and 6,328 outliers were excluded (Figure 1). From the 87,361 valid measurements, hourly averages were computed. Among the 3,230 hourly PM_2.5_ concentration measurements, the median PM_2.5_ concentrations were 164.2 µg/m^3^ [IQR 107.0–272.2] with a minimum of 31.5 and a maximum of 5,525.7 µg/m^3^. The median 24-hour exposure per child was 144.2 µg/m^3^ [118.4–241.0], with a minimum of 39.8 and a maximum of 696.3 µg/m^3^. Children from G2 were exposed to the highest PM_2.5_ concentrations with a median of 270.2 µg/m^3^ compared to 134.73 in G1 and 142.4 in G3 (p = 0.016) (Table 4).

**Table 4:** PM_2.5_ concentrations (µg/m^3^) according to children’s characteristics.

| Variable | Modality | N | Median [Q1–Q3] | Mean (SD) | Min – Max | P-value* |
| --- | --- | --- | --- | --- | --- | --- |
| Hourly PM<sub>2.5</sub> (all measurements) |  | 3230 | 164.2 [107.0 – 272.2] | 238.9 (322.8) | 31.5 – 5525.7 |  |
| 24-hour PM<sub>2.5</sub> exposure per children |  | 29 | 144.2 [118.4 – 241.0] | 250.0 (135.8) | 39.8 – 696.3 |  |
| Gender | Female | 2177 | 153.2 [105.2 – 258.5] | 241.1 (358.5) | 33.5 – 5525.7 | 0.299 |
|  | Male | 1053 | 194.1 [112.3 – 290.5] | 234.56 (232.1) | 31.5 – 3041.8 |  |
| Schoolchildren | 0 | 358 | 233.9 [103.8 – 373.2] | 413.3 (652.6) | 34.6 – 3747.2 | 0.188 |
|  | 1 | 2872 | 158.7 [107.6 – 259.2] | 217.22 (244.9) | 31.5 – 5525.7 |  |
| Asthma | 0 | 2768 | 170.2 [105.9 – 281.9] | 248.84 (344.0) | 31.5 – 5525.7 | 0.389 |
|  | 1 | 462 | 148.3 [111.8 – 212.7] | 179.72 (123.4) | 36.4 – 1597.8 |  |
| Study Group | G1 | 1437 | 134.7 [105.8 – 192.0] | 167.21 (135.7) | 34.1 – 3315.4 | 0.016<sup>1</sup> |
|  | G2 | 820 | 270.2 [203.6 – 353.1] | 385.4 (527.0) | 31.5 – 5525.7 |  |
|  | G3 | 973 | 142.4 [85.0 – 275.4] | 221.5 (242.8) | 34.1 – 2084.3 |  |

\* Generalized linear mixed-effects model with repeated measures
<sup>1</sup> Post-hoc test : G1 vs G2 ( $\beta = -457.0$ , $p = 0.018$ ), G2 vs G3 ( $\beta = 415.2$ , $p = 0.034$ ), G1 vs G3 ( $p = 0.957$ ).

The average hourly PM_2_._5_ concentrations varied throughout the day, with peak exposures around 7 a.m., 2 p.m., and 7 p.m. and concentrations well above WHO recommendations, overall (Figure 2A). Group 2 had the highest PM_2.5_ exposure levels, with a clear peak between 12 p.m. and 3 p.m., reaching over 750 µg/m^3^, while groups 1 and 3 had lower concentrations, slightly higher in group 3 compared to group 1 (Figure 2B). When stratified by gender, only Group 2 showed higher exposure among girls than boys during the day (Figure 2C.). Similarly, when stratified by school status, unschooled children in group 2 were exposed to significantly higher levels than their schooled counterparts, with hourly peak concentrations exceeding 2000 µg/m^3^ between 12 noon and 3 p.m., a difference not observed in group 3 (Figure 2C.).

**Figure 2.**
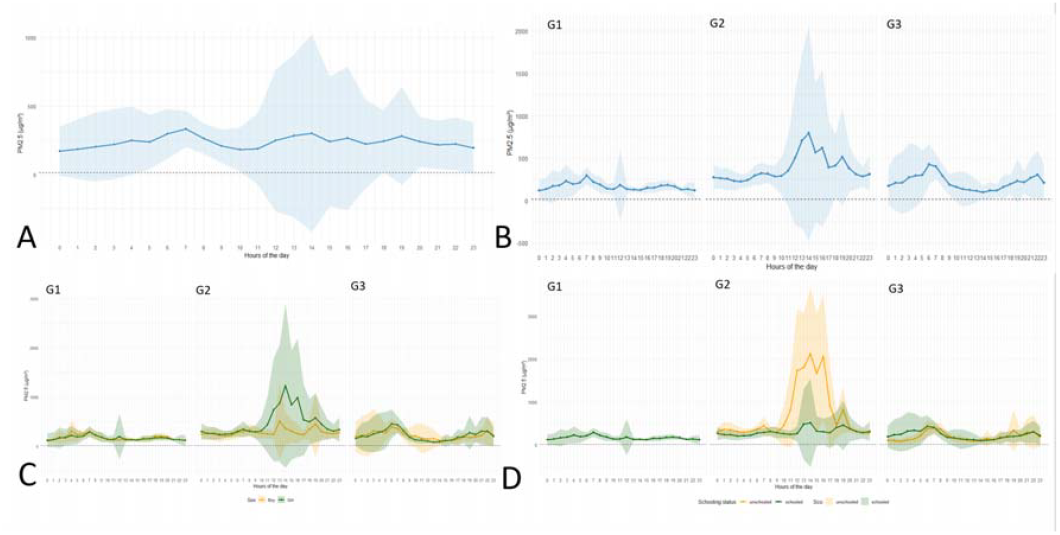
Average hourly concentration of PM_2_._5_ over 24 hours according to various characteristics (N=29) **A**. Mean hourly PM_2_._5_ concentration over a 24-hour period, **B**. Mean hourly PM_2_._5_ concentration over 24 hours by group **C**. Mean hourly PM_2_._5_ concentration over 24 hours by group and sex, **D**. Mean hourly PM_2_._5_ concentration over 24 hours by group and schooling status. The black dashed line represents the WHO air quality guideline threshold not to be exceeded: 15 µg/m^3^ over 24 hours.

When analyzing the association between PM_2.5_ exposure and ppFEV_1_, we evaluated concentrations over three time windows prior to spirometry: the 24-hour average, the maximum 1-hour, and the maximum 8-hour values. No significant associations were observed at any of these intervals. Specifically, a increase of 1 µg/m^3^ in average PM_2_._5_ concentration during the 24 hours preceding the spirometry measurement was not associated with a decrease in ppFEV_1_ (p> 0.05) (Table 5).

**Table 5.** Association between short-term PM_2_._5_ exposure and ppFEV_1_.

| PM<sub>2.5</sub> exposure | Estimate ($\beta$) | 95% CI | p-value |
| --- | --- | --- | --- |
| Lag mean 24 (mean over 24h before FEV<sub>1</sub>) | 0.011 | [-0.0047 - 0.0267] | 0.162 |
| Max 1 hour in the last 24 hours before FEV<sub>1</sub>) | 0.002 | [-0.0025 - 0.0055] | 0.478 |
| Max 8-hour moving average (last 24 hours) | 0.004 | [-0.0036 - 0.0124] | 0.286 |

\*Models adjusted for sex, school, group, and asthma-related spirometry, Estimates correspond to the change in ppFEV<sub>1</sub> per 1 $\mu\text{g}/\text{m}^3$ increase in PM<sub>2.5</sub>, Random intercepts by individual (Code), with AR(1) correlation structure over time, Imputation on PM<sub>2.5</sub> interpolated by child based on valid values, respecting chronology and without imputing large data gaps (>6h).

The results of the mixed-effects distributed lag polynomial analysis showed that exposure to PM_2.5_ was associated with a transient decrease in ppFEV_1_, mainly between 0 and 4 hours post-exposure. Significant effects were observed between the 0 and 2^nd^ hours, reaching an estimated minimum at Lag 0 hours (β = −2.21; CI [−3.74; −0.69]). A moderate, trans improvement in ppFEV_1_ was then observed between 5 and 16 hours, peaking around Lag 10 (β = 0.68; CI [0.08; 1.27]), followed by a further non significant decline between lag 16 to 36 and 42 to 48 hours (Figure 3A.). Models stratified by study group showed distinct lag-response profiles: Group 2 had a broader but non-significant variation, while Group 3 closely matched the overall pattern (Figure 3A.). Time-of-day stratification revealed, non-significant curves (Figure 3C.).

**Figure 3.**
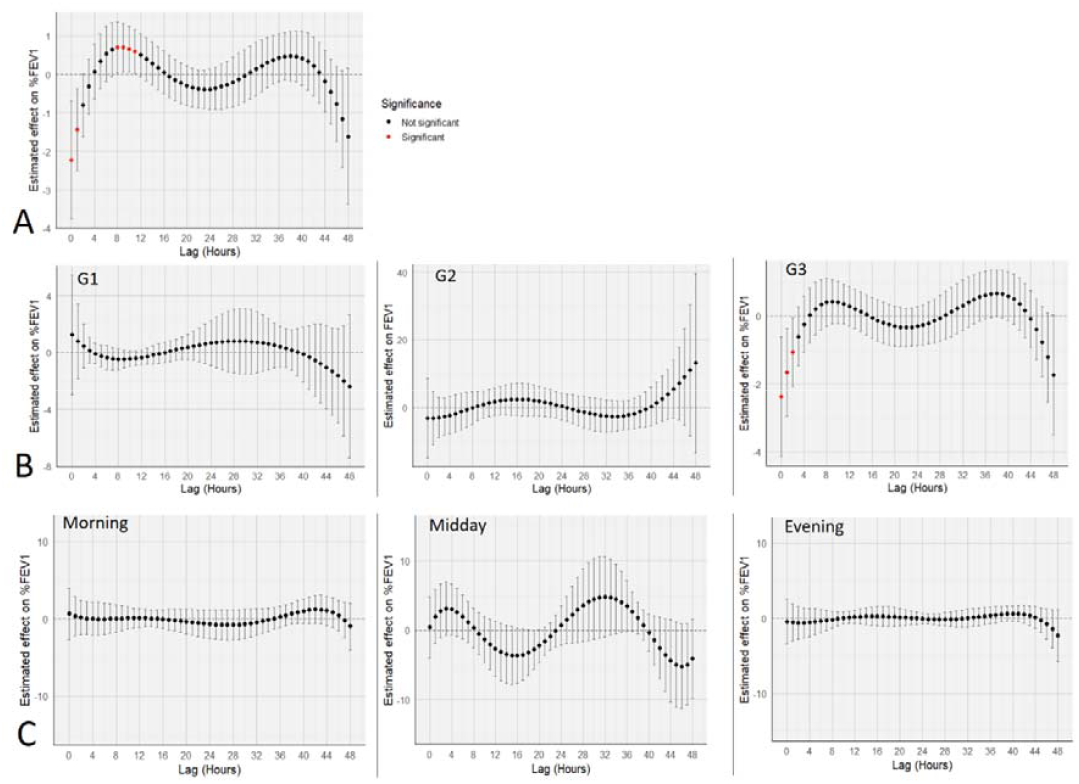
Estimated lag effect of personal hourly PM_2_._5_ on ppFEV_1_ A. Hourly delayed effect of personal PM_2_._5_ on ppFEV_1_ (n = 29). Estimates from a mixed-effects distributed lag polynomial model (5th degree, AR(1)), adjusted for gender, asthma, and education. Variation associated with an IQR of 120.9 µg/m^3^. B. Hourly delayed effect of PM_2_._5_ by group (G1, G2, G3). Same model as A; IQR = 49.7 µg/m^3^, for G1, 180.9 µg/m^3^ and 96.5 µg/m^3^ for G3. C. Hourly delayed effect of PM_2_._5_ according to time of day (morning, midday, evening). Same model and adjustments as A; IQR = 120.9 µg/m^3^.

## DISCUSSION

Our study shows that collecting individual respiratory function data using portable spirometry and measuring personal PM2.5 exposure with mobile sensors is feasible among children living in precarious urban settings in Abidjan, Côte d’Ivoire. Substantial intra-day variability was observed, with more pronounced lung function impairment in the morning. PM2.5 exposure levels were markedly elevated, particularly among children whose mothers smoked fish with inter-individual variability, partly influenced by gender and school attendance. Notably, we identified a short-term decline in lung function within hours of PM_2.5_ exposure, highlighting critical windows of respiratory vulnerability.

A high proportion of volume flow curves was collected, reflecting good engagement and feasibility of repeated ambulatory spirometry on children. However, fewer than a third of the curves were valid, highlighting technical limitations particularly the absence of feedback or motivational features in the devices used. Previous studies have shown that child-friendly interactive tools can improve spirometry quality (26). At inclusion, prevalence of asthma respiratory symptoms was relatively low, particularly for wheezing in the past 12 months (6.9%), although nocturnal dry cough was more commun. These findings remain consistent with those reported in similar African settings (14,27,28). Despite limited respiratory symptoms, baseline spirometry revealed significant lung function impairment, with 13% of children showing presumptive asthma. Reduced ppFEV_1_ was observed in several participants at baseline and became more frequent during ambulatory follow-up, with a similar trend for FVC across subgroups, suggesting ongoing functional decline. Part of this decline may be related to suboptimal efforts during unsupervised testing, which is known to yield lower and more variable FEV_1_ and FVC values compared to supervised assessments (29). Nevertheless, ambulatory spirometry results were consistent with baseline measurements, reinforcing the reliability of our data. A substantial number of children exhibited abnormal spirometric patterns despite infrequent and mild symptoms. This discrepancy highlights the potential for chronic air pollution exposure to cause subclinical respiratory impairment that may not be captured through symptom-based assessments. Our findings support the hypothesis that long-term exposure to fine particulate matter contributes to reduced lung function in children, even in the absence of overt clinical symptoms or diagnosed asthma (1,6). Children exposed to charcoal (G1) consistently showed lower FEV_1_ and FVC values at baseline and during ambulatory spirometry suggesting a possible link between long-term indoor air pollution and reduced expiratory airflow, consistent with prior studie (30).

Personal ambulatory PM_2_._5_ monitoring was successfully implemented, capturing almost 2/3 of expected data confirming its feasibility in children from vulnerable urban areas. Median 24-hour PM_2_._5_ per child concentrations reached nearly ten times above WHO guidelines with high variability across sex, schooling, exposure group, and time of day, reflecting consistently elevated exposure levels among children living in these neighborhoods in Abidjan, as supported by previous studies using fixed-site monitoring (14,28). The elevated exposures in G2 notably among unschooled girls are likely attributable to their involvement in maternal fish smoking activities. A previous field survey revealed that 79.1% of helpers in smoking sites in the city of Katiola, Côte d’Ivoire were minors, highlighting that frequent child involvement may substantially increase their exposure to air pollutants (31). In contrast, the type of domestic fuel (charcoal vs. gaz) was not significantly associated with exposure levels suggesting that children’s daily routines and time spent in polluted microenvironments may play a more important role than household fuel use.

Linking ppFEV_1_ with PM_2_._5_ exposure revealed a transient decline in lung function, with the strongest effect observed few hours after an increase in PM_2_._5_ levels equivalent to the interquartile range of daily exposure. These findings align with prior studies reporting reduced FEV_1_ following short-term PM_2_._5_ exposure (1,10). Most previous studies relied on fixed-site monitors, whereas our use of personal sensors may better reflect individual exposures (32). Variability in lag effects may relate to underlying inflammatory responses that precede measurable functional decline, as previously suggested (2). Our cohort’s substantially higher exposures likely contributed to the observed effects, even in a predominantly non-asthmatic population. In our cohort, morning FEV_1_ values were systematically lower, in line with the known circadian variation in lung function among children. This may amplify the effect of early-day exposure peaks, particularly in settings with elevated ambient PM_2.5_.

This study has several limitations. The small sample size and short duration (seven days) limit the ability to capture longe-term variability. A limited number of valid spirometry curves may have reduced statistical power and representativeness. Environmental factors such as temperature and humidity, which can influence lung function, were not available for adjustment (33,34). The focus on PM_2_._5_ alone, without accounting for co-pollutants, restricts assessment of co-exposure effects. Additionally, respiratory symptoms were not recorded during the monitoring period due to the complexity of the symptom diary. Future studies could benefit from simplified digital tools to enhance adherence and capture symptom data more effectively (35). Despite these limitations, this study provides novel, context-specific insights into real-time and short-term effects of PM2.5 exposure on lung function in children living in under-resourced urban settings.

To our knowledge, our study is among the first to combine repeated ambulatory spirometry with personal PM_2_._5_ monitoring in this context. The variability observed in both lung function and exposure, supports the relevance of individualized, mobile monitoring approaches over fixed-site monitoring. The respiratory impacts associated with extreme exposures, particularly in settings involving fish smoking and charcoal use, underline the need for researches and interventions that account not only for household fuels, but also for daily behaviors and time spent in polluted micro-environments since birth. Moving forward, research should focus on identifying high-risk environments and activities based on children’s age and routines, in order to better inform targeted public health strategies.

## ACKNOWLEDGEMENTS

The authors would like to thank the study participants, including the children and their families, for their valuable participation, as well as the students who contributed to data collection and analysis. They express their gratitude to AQuiRespi, a collaborative platform of the French Regional Health Agency (ARS) dedicated to respiratory health research and innovation, for their interest in the topic and support. They also thank the PAC-CI Research Center (ANRS site) in Abidjan for their logistical and technical support in implementing the study. Finally, we would like to thank the APIMAMA project, funded by the ANR, for its support in the implementation of this project.

## AUTHOR CONTRIBUTORS

AP wrote the study protocol, coordinated study implementation, contributed to conceptualization, data curation, methodology, and writing, collected data, trained children on using the spirometry devices, and drafted the first version of the manuscript. MY, SA, and SD were involved in data collection. CL, MD, FT, MF, and VY contributed to conceptualization, methodology, supervision, and validation of the manuscript. SG contributed to conceptualization and methodology. OM designed the study, supervised its scientific implementation and analysis, and contributed to manuscript write up and validation. All authors reviewed the manuscript and approved the final version.

## DATA AVAILABILITY

Study data will not be publicly available. Data could be made available by study group to any researcher interested. Deidentified participant data and a data dictionary can be made available and shared under a data transfer agreement. Requests for access to the APIMAMA Kids study data should be sent to.

