## Appendix 1 for "Feasibility and results of joint ambulatory monitoring of exposure to particulate matter pollution and lung function in children in Abidjan, Côte d’Ivoire, a cross-sectional observational study"

Appendix1 – Comparison of medians of the number of flow-volume curves obtained in children

| Variable | Modality | Number | Median [Q1-Q3] | P-value* |
| --- | --- | --- | --- | --- |
| Overall |  | 29 | 36 [29-42] | -- |
| Sex | F | 18 | 33.0 [27.2-36] | **0.005** |
|  | M | 11 | 42.0 [36.5-59] |  |
| Asthma | 0 | 25 | 36.0 | 0.825 |
|  | 1 | 4 | 35.0 [32-43] |  |
| Schoolchildren | 0 | 3 | 32.0 [29.5-46.5] | 0.830 |
|  | 1 | 26 | 36.0 [30-41.8] |  |
| Study group | G | 10 | 34.0 [30-36] | 0.071 |
|  | G2 | 9 | 33.0 [25-42] |  |
|  | G3 | 10 | 41.5 [36.5-59.5] |  |
| *Wilcoxon and Kruskal-Wallis test | | | | |
